# Advanced analysis in epidemiological modeling: Detection of wave

**DOI:** 10.1101/2021.09.02.21263016

**Authors:** Abdon Atangana, Seda İğret Araz

**Affiliations:** Institute for Groundwater Studies, Faculty of Natural and Agricultural Sciences, University of the Free State, South Africa; Department of Medical Research, China Medical University Hospital, China Medical University, Taichung, Taiwan; Siirt University, Department of Mathematics Education, Siirt, Turkey

**Keywords:** Strength number, second derivative analysis, waves, piecewise modeling

## Abstract

Some mathematical concepts have been used in the last decades to predict the behavior of spread of infectious diseases. Among them, the reproductive number concept has been used in several published papers for study the stability of the spread. Some conditions were suggested to predict there would be either stability or instability. An analysis was also suggested to determine conditions under which infectious classes will increase or die out. Some authors pointed out limitations of the reproductive number, as they presented its inability to fairly help understand the spread patterns. The concept of strength number and analysis of second derivatives of the mathematical models were suggested as additional tools to help detect waves. In this paper, we aim at applying these additional analyses in a simple model to predict the future.

## 1 Introduction

Mathematical models of spread of disease have been used successfully with limitation to predict future behaviors of the spread. The main aim is to have an asymptotic idea of what the spread will look like in a near future, such that measures could be taking to help control and eradicate the spread of the virus. Several mathematical concepts have been suggested to help better analyze these models. During the modelling process, mathematicians or modelers first translate the observed facts into mathematical equations using either ordinary differential equations or partial differential equations [1–4]. The obtained system of equation either linear or nonlinear is further analyze. The first analysis consisting in obtaining equilibrium points (disease free and endemic). This analysis is often followed by analysis of stability that include global and asymptotic global stability, conditions under which the infectious classes will decrease, increase, or stay constant. The reproductive number that can be obtained in different way for instance using the next generation of matrix is obtained to help predict either the spread will be stable or not. Then finally the study of the sign for the first derivative of the Lyapunov function associate to the system to also give clear idea of the stability. In thousands of papers published in this topics, similar analysis can be found, so far there is not major contribution that have been done after the idea of reproductive number was suggested. However, while checking with great care predictions done while using this analysis, one will easily see that this analysis cannot help humans predict waves in spread. For example, several mathematical models have been suggested to predict the spread of Covid-19, unwillingly no analysis help to predict the numbers of waves that humans will faced by Covid-19 spread [5–12]. In fact, some researchers showed that, the reproductive number was not sufficient to provide a clear behavior of the spread, although no attention was paid to such analysis, one will agree that suggested mathematical models and analysis done in the last decades have not really help to predict waves. Very recently, additional sets of analysis were suggested, first an evaluation equilibrium point of second derivative of the model, evaluation of conditions under which the second derivatives of infections classes are positive, negative or zero, calculation of strength number using next generation matrix and evaluation of sign of second derivative of Lyapunov function associate to the model. Additionally, it was also suggested that model should be derived with piecewise derivatives, in this paper we will apply that analysis to a simple epidemiological model.

## 2 A SEIR model

While the SEIR model has been used and analysed in many research works to predict spread of some infectious diseases. This model has been used to calculate the so-called reproductive, also they have calculated its respective equilibrium points including disease-free and endemic points. However, the model has not been used to check either the model can predict waves, or if the equilibrium points are local maximum or local minimum. In this section, a SEIR model will subjected to a different analysis that may help if such model is suitable for describing spread with waves.

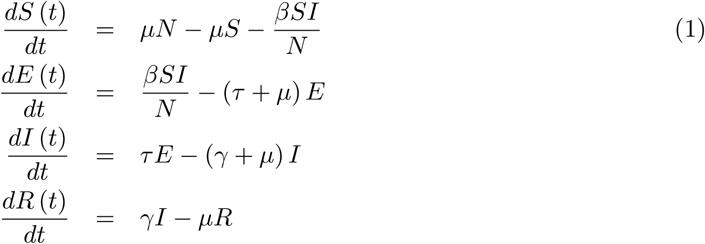

where *N* = *S* + *E* + *I* + *R*. The initial conditions are as follows;

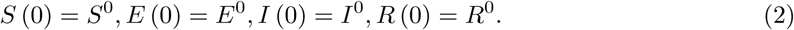

Here *S* (*t*) is the class of the susceptible individuals. *E* (*t*) is the class of the exposed individuals. *I* (*t*) is the class of the infected individuals. *R* (*t*) is the class of the recovered individuals.

### 2.1 Positiveness and boundness of solutions

Just to start with primaries analysis, at least to show that the solutions are positive since they depict real world problem with positive values. In this subsection, we examine the conditions under which the positivity of the solutions of the considered model are satisfied. Let us start with the class *E* (*t*)

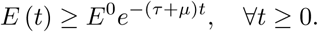

For the function *I* (*t*), we have the following inequalities

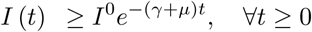

and

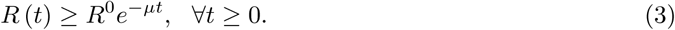

We shall define the norm

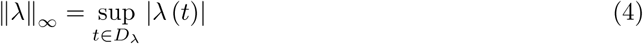

where *D*_*λ*_ is the domain of *λ*. Using the above norm, we get the following inequality for the function *S* (*t*),

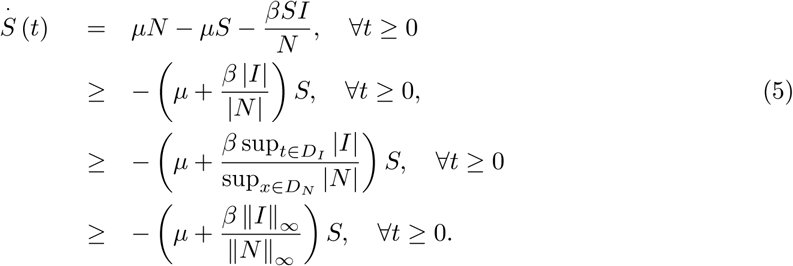

This yields

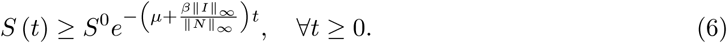

### 2.2 Analysis of equilibrium points

In this subsection, we give a detailed analysis about equilibrium points. Disease-free equilibrium for this model is

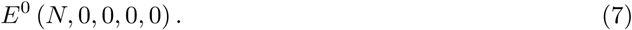

To get the endemic equilibrium points, we need to solve the following system

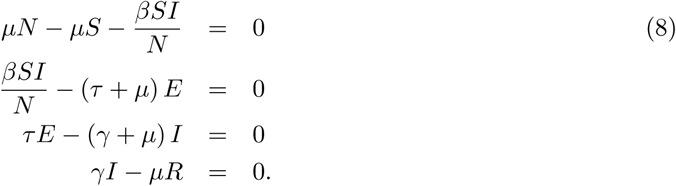

Then the endemic equilibrium points are

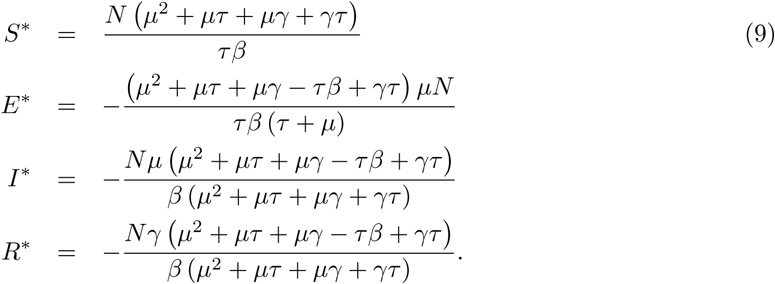

The above endemic eqilibrium are valid if the following inequality holds

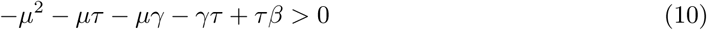

namely

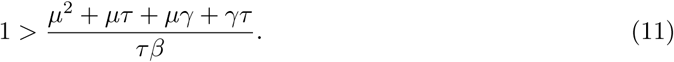

### 2.3 Reproduction number

To proceed with our preliminary analysis, we shall present here the derivation of the so-called reproductive number. This is important value in the field of epidemiological modelling, as it was revealed to help to have an understanding the stability conditions. It was documented that, if this number is less than 1, we can expect stability, however if this number is greater than 1, we could expect instability. Nonetheless, it was reported that the value is obtained with different ways. However, in this case we will utilize the next matrix approach to achieve the reproductive value.

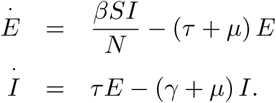

Using the next generation matrix approach [13], we calculate the matrices *F* and *V* ^-1^ as

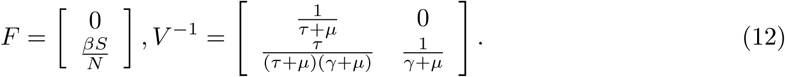

Then, the reproduction number is obtained as

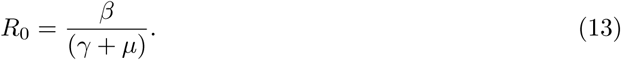

### 2.4 Strength number

Although the so-called reproductive number has been used in thousands of research papers in epidemiological modelling with some success and great limitations. While this concept has been employed to determine either or not the spread will be severe, some great weaknesses have been pointed out by some researchers, for example they sadly realized that such value is not unique as it can be obtained via different methods. Another issue was raised that, this number should have been a function of time not a constant value. Additionally, it was also noticed that a reproductive number cannot be used to indicate either a model will predict waves or not. Very recently an alternative number was suggested and was called strength number, of course this number will be subjected to several test to see either it can be used to help to detect some complexities in the spread, at least it this number can help detect waves in a spread. The value was derived using the next generation matrix, by taking the second derivative of infectious classes. In this section, we will present the strength number associate to SEIR model.

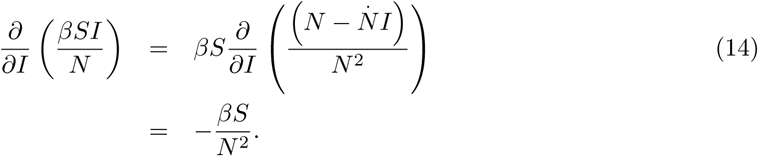

In this case, we can have the following

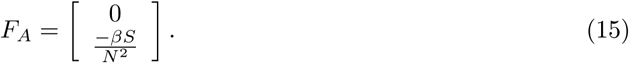

Then

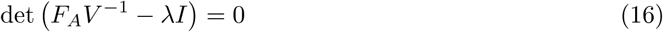

yields

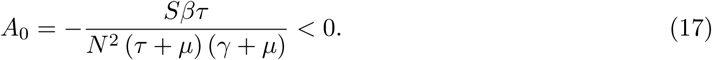

Having obtained the strength number to be less than zero will lead us to a great conclusion that will further be confirmed using a different analysis consisting in finding local minimum, local maximum, and inflection points. Thus, having strength a negative number is an indication that the SEIR model will have a single magnitude, either maximum point with two inflection points indicating a single wave or infection will decrease rapidly from the disease-free equilibrium and with renewal process the infection will raise after a minimum point and then be stabilized or stop latter on. This will be confirmed while studying sign of second derivative of infectious classes.

### 2.5 First derivative of Lyapunov

For the endemic Lyapunov function, {*S, E, I, R*} ,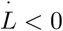 is the endemic equilibrium *E*^*^.

#### Theorem 1.

When the reproductive number *R*_0_ *>* l, the endemic equilibrium points *E*^*^ of the SEIR model is globally asymptotically stable.

**Proof**. For proof, the Lyapunov function can be written as

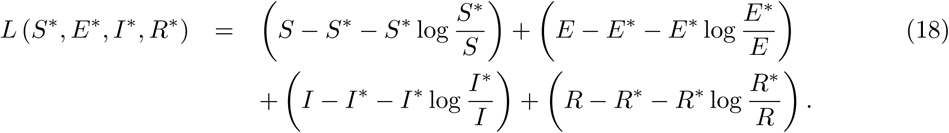

Therefore, applying the derivative respect to *t* on both sides yields

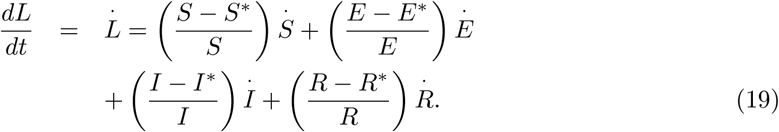

Now, we can write their values for derivatives as follows

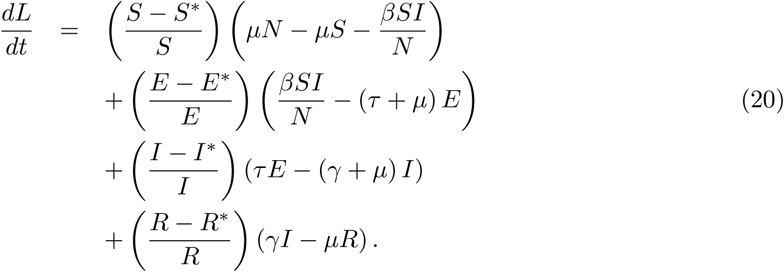

Putting *S* = *S* − *S*^*^, *E* = *E* − *E*^*^, *I* = *I* − *I*^*^, *R* = *R* − *R*^*^ leads to

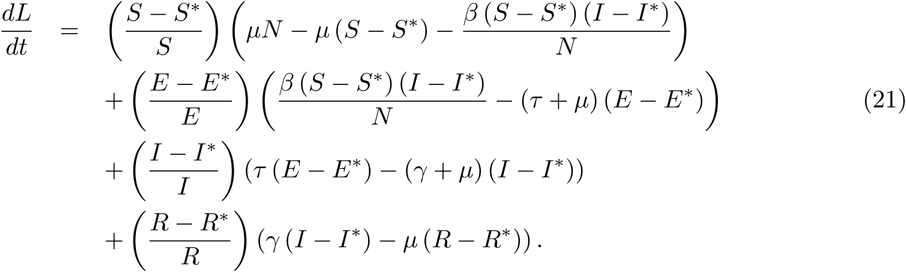

We can organize the above as follows

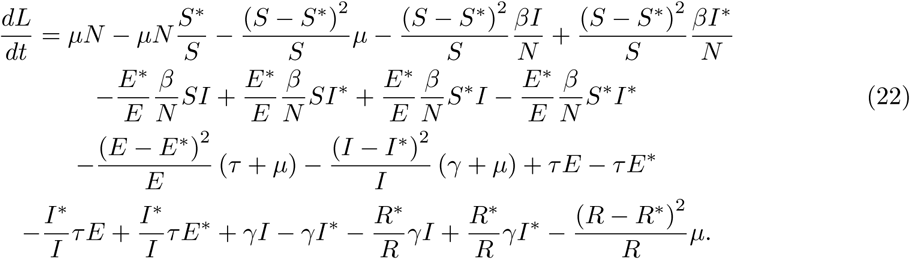

To avoid the complexity, the above can be written as

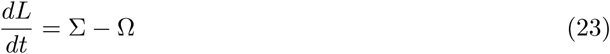

where

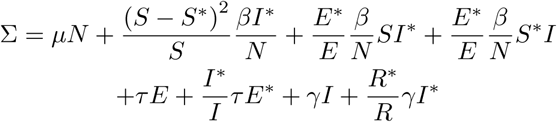

and

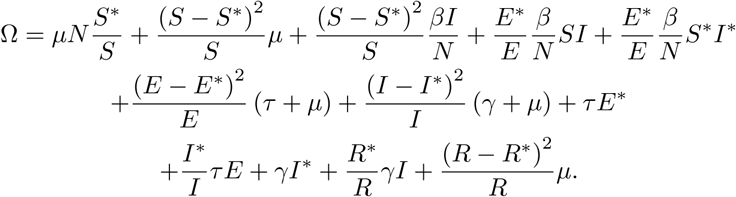

It is concluded that if ∑ < Ω, this yields 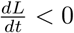, however when *S* = *S*^*^, *E* = *E*^*^, *I* = *I*^*^, *R* = *R*^*^

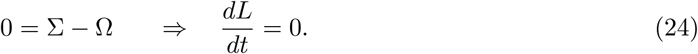

We can see that the largest compact invariant set for the suggested model model in

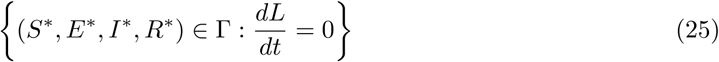

is the point {*E*_*_} the endemic equilibrium of the considered model. By the help of the Lasalle’s invariance concept, it follows that *E*_*_ is globally asymptotically stable in Γ if ∑ < Ω.

### 2.6 Second derivative of Lyapunov

While the sign of the first derivative of a Lyapunov function helps to appreciate the stability, one should note that, analysis of first derivative of a given function cannot fully help to understand the variabilities of the function under study. Further analysis are required to all the particularities of the variations. We thus present an analysis of second derivative of the associated Lyapunov function of our model.

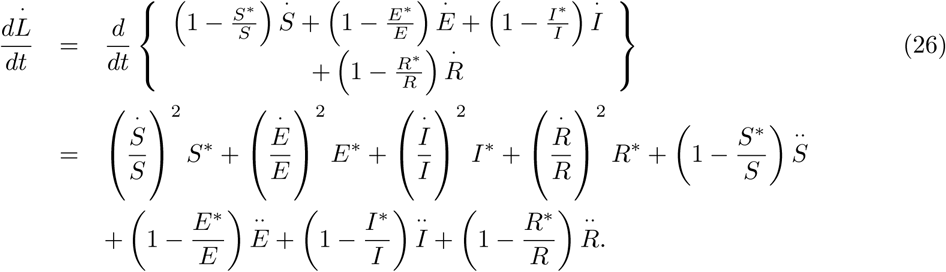

Here

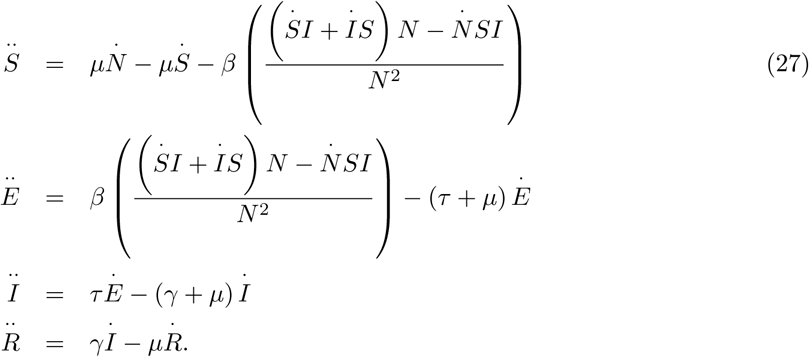

Then, we have

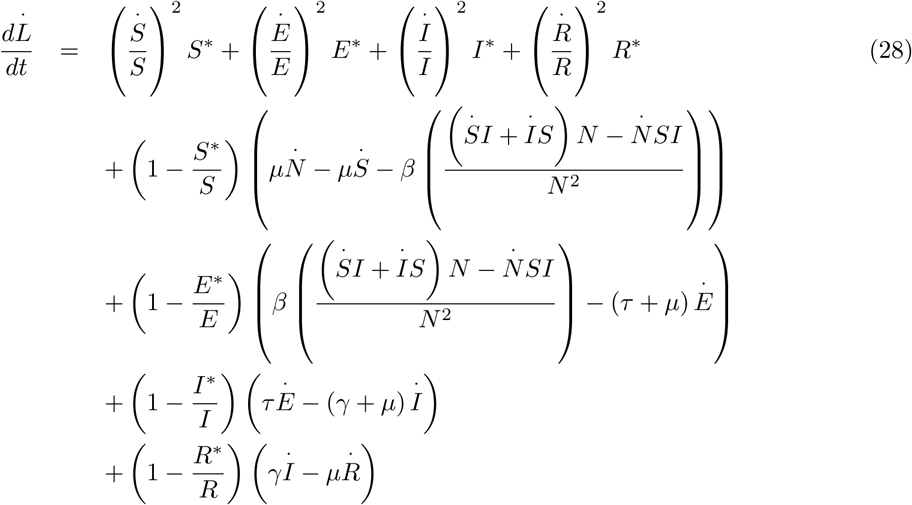

and

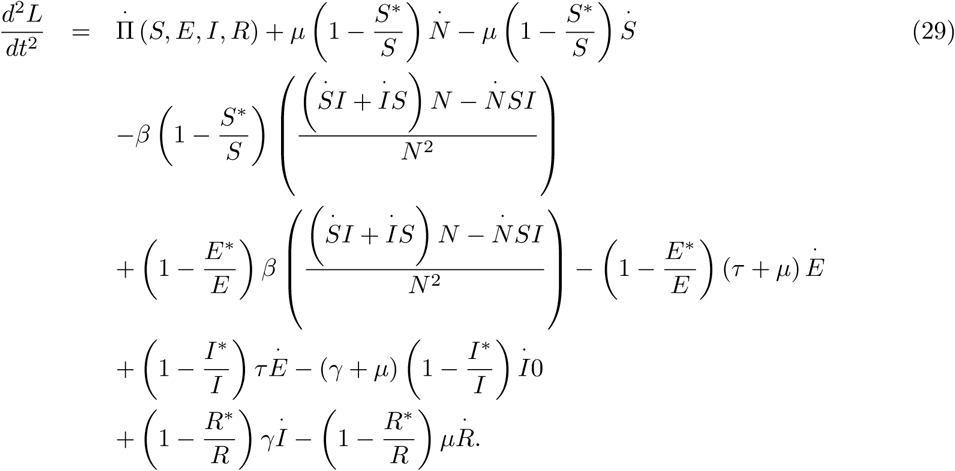

After replacing 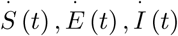 and 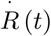 by their formula from the considered model and putting all together, we can get

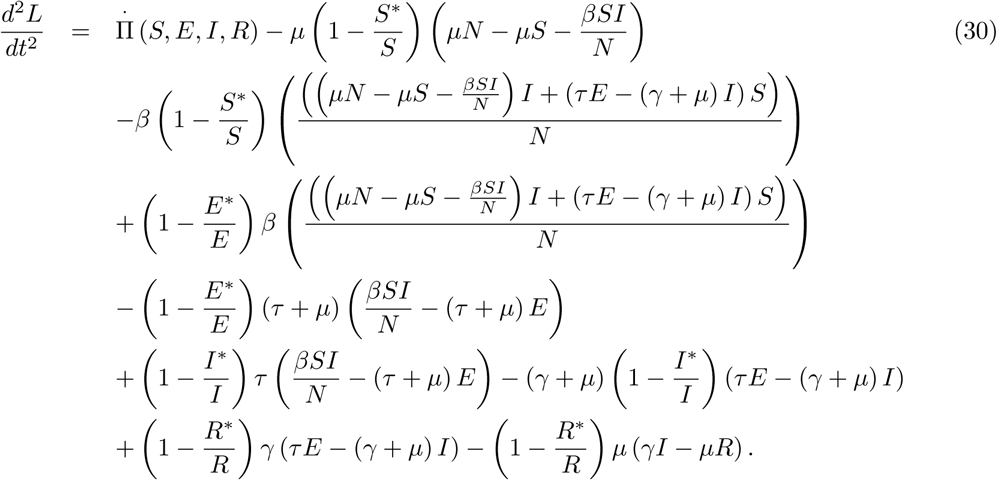

For simplicity, we can write

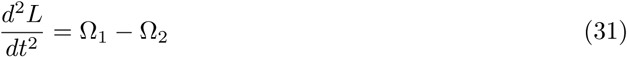

where

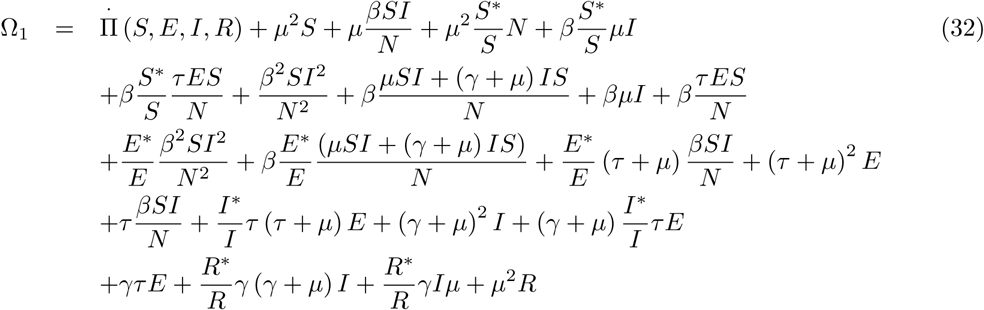

and

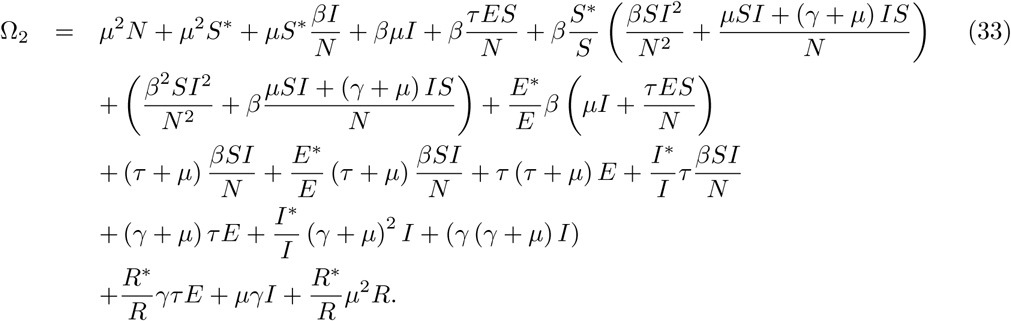

It can be seen that

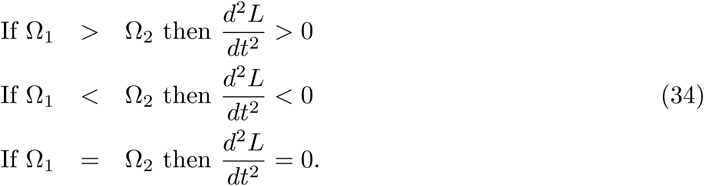

### 2.7 Equilibrium points for second order

Since the equilibrium points also help to get information on curvatures, in this subsection, we aim to find the equilibrium points of second order derivative of our solutions. Thus, we write

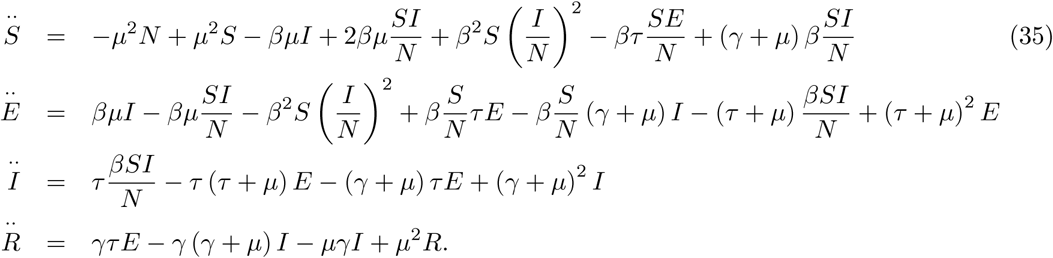

The disease-free equilibrium point of second order derivative of model is

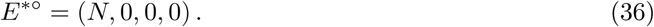

and the endemic equilibrium points of second order derivative of the model is

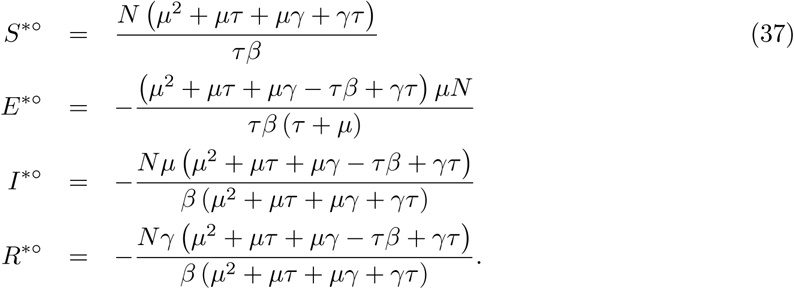

For the classes *I* (*t*), *E* (*t*) to increase, we need

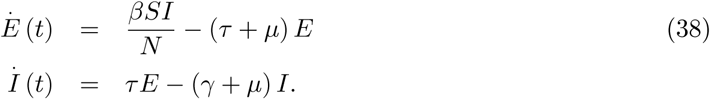

Indeed for the endemic process, we can consider the following

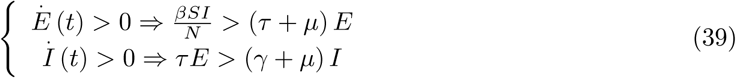

and since 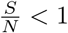

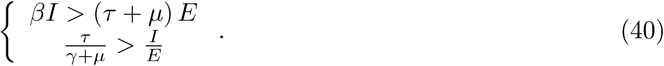

Thus,

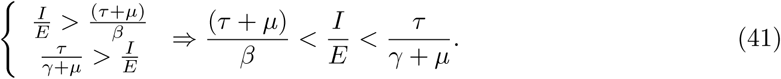

*E* (*t*) and *I* (*t*) classes will increase if the following conditions

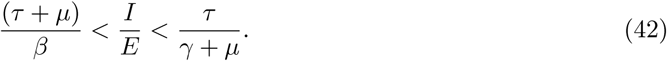

We are n01w checking local maximum, local minimum or inflection points. We have two equilibrium point including the disease-free and endemic. To achieve this, we consider the second derivative of the infectious classes

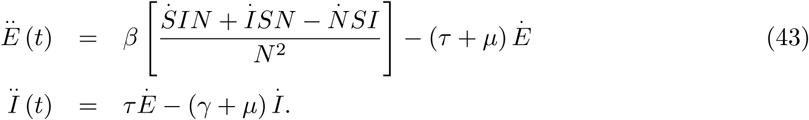

At *E*^0^, we have that

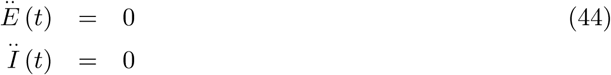

which can be considered as a changing point of the spread. Meaning at this point, we could see any increase in number of infectious classes.Also, at *E*^0^*, endemic equilibrium 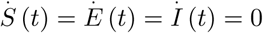. Thus *Ë* (*t*) = *Ï* (*t*) = 0 also. The model predicts two changing points, therefore, it is expected to have either a maximum or minimum point.

We shall evaluate the sign of *Ë* and *Ï* functions

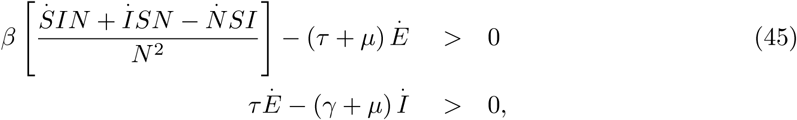

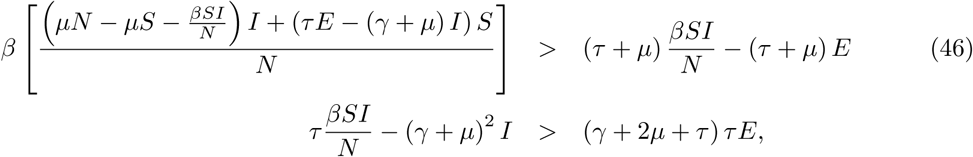

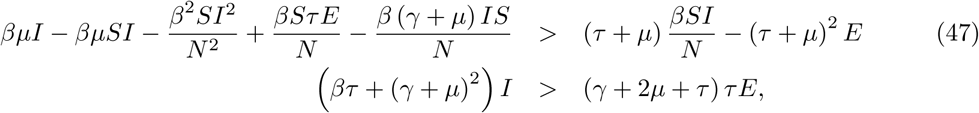

and

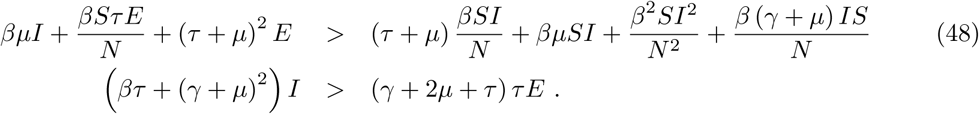

Since 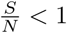, then we can write

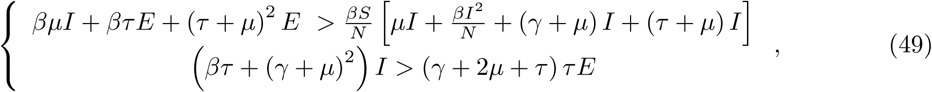

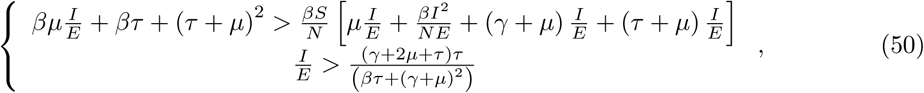

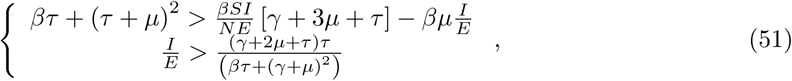

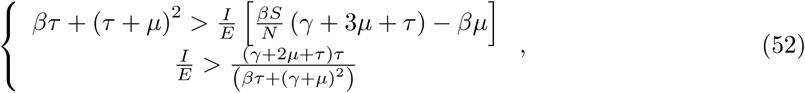

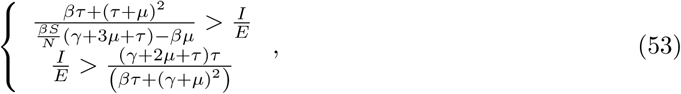

with 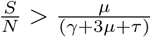. Thus we have the following condition

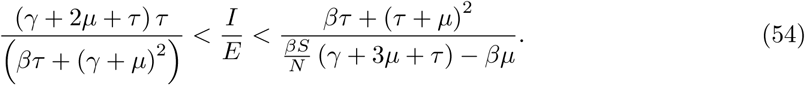

On the other hand if

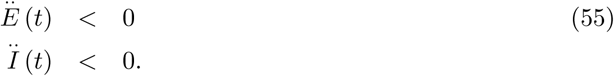

From this inequality, we have

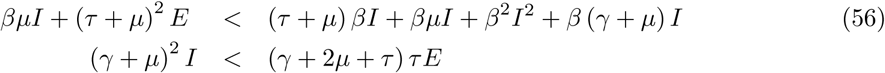

and

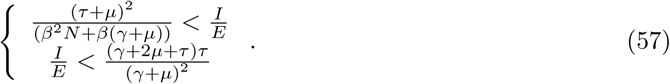

Therefore

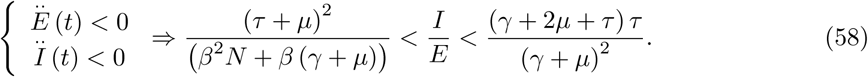

In Figure 1, we present a numerical simulation showing the condition under which the two infection classses admit negative second derivatives. The theoretical interpretation of such condition will therefore follow;

**Figure 1.**
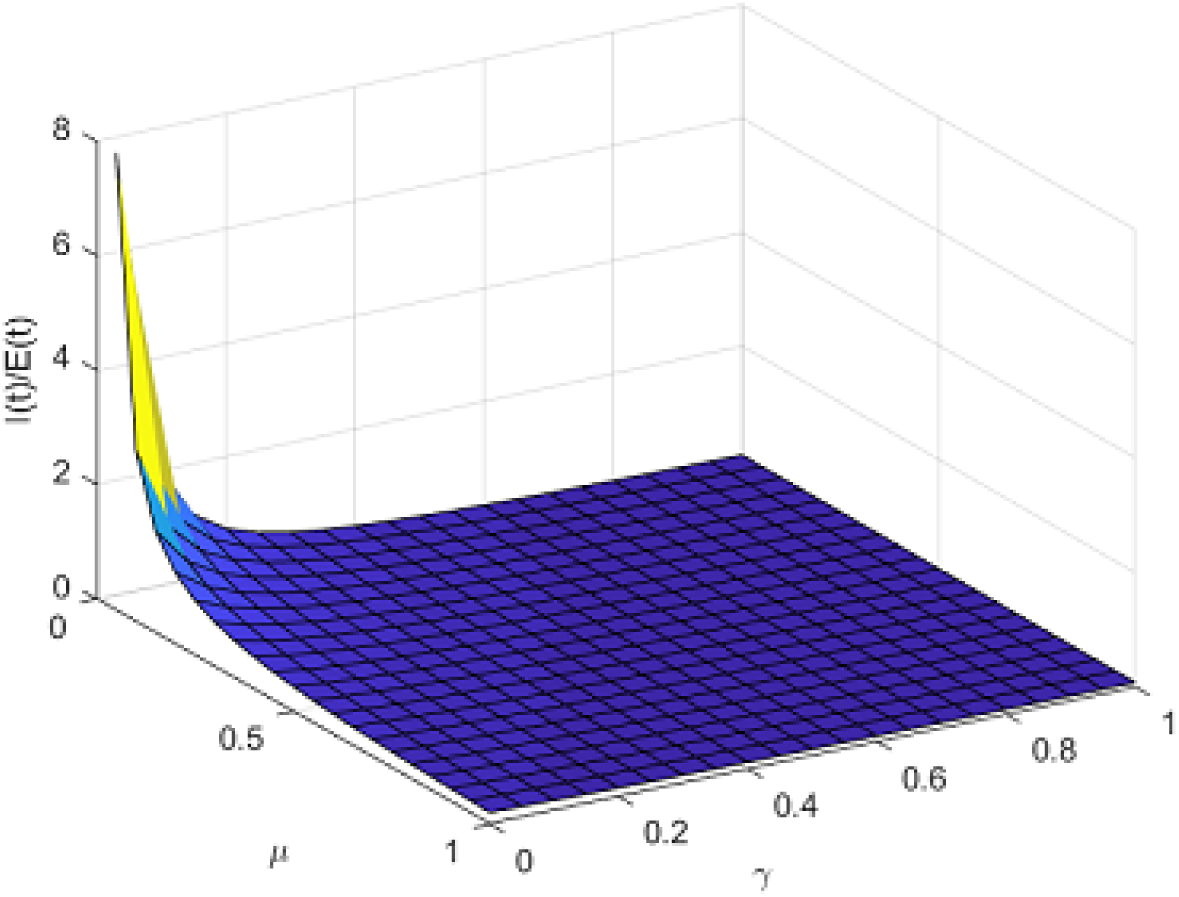
Waves test condition.

Under this condition, the SEIR model may not be able to depict more than two waves. The model shows that the spread will have only one peak and then die out later with no renewal force. In literature, however such model has been used to depict spread of infectious diseases with different waves.

## 3 Numerical solution of the model with exponential decay process

In this section, we present a numerical scheme based on Newton polynomial for the numerical solution of the considered model with piecewise differential and integral operators [14,15]. We start with the piecewise SEIR model with classical and stochastic Caputo-Fabrizio case which is given by

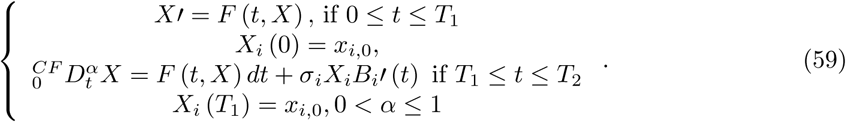

Here

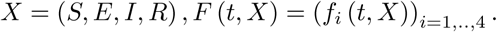

Applying the associated integral, we can have

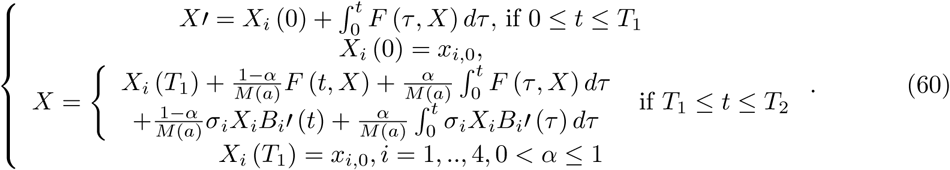

We divide [0, *T*] in two

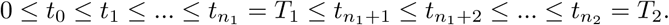

Interpolating *f*_*i*_ (*t, X*_*i*_) using the Newton polynomial within [*t*_*n*_, *t*_*n*+1_] yields

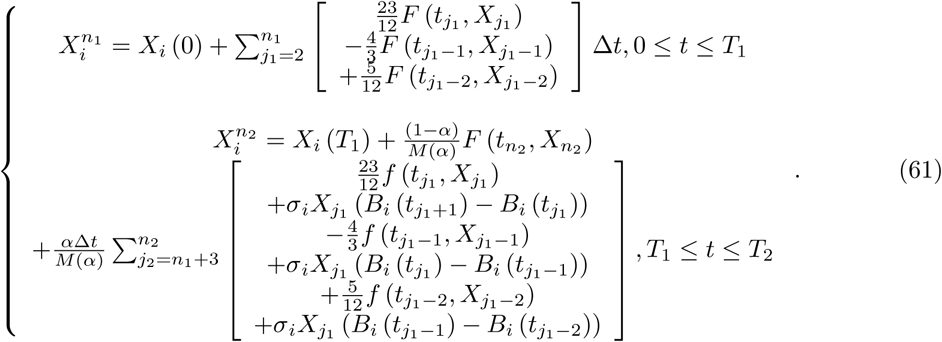

The parameters and initial conditions are given as

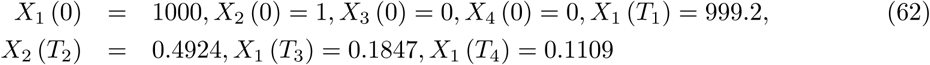

and

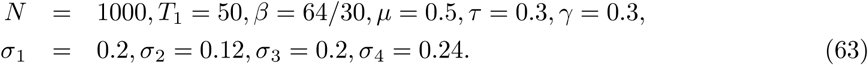

The simulation of the numerical solution of the model is depicted in Figure 2.

**Figure 2.**
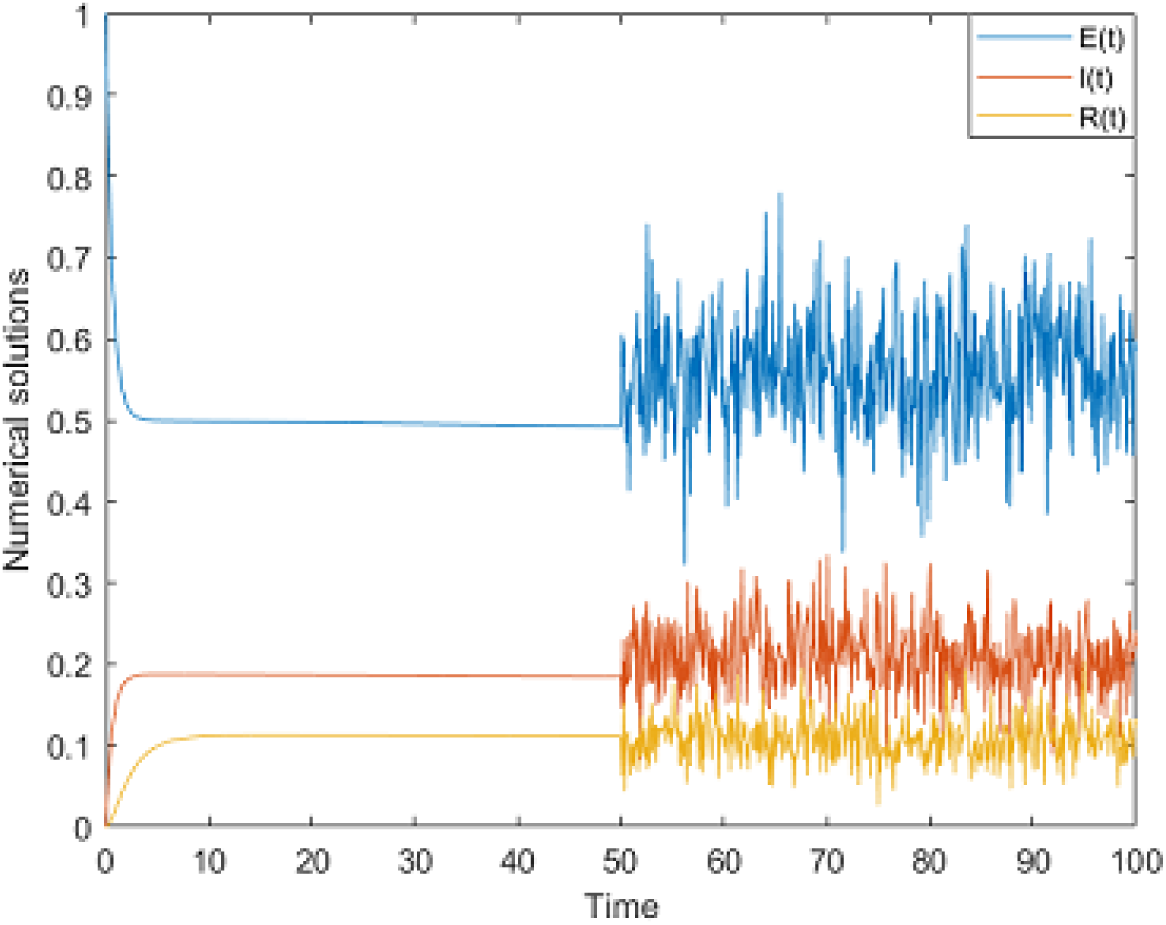
Numerical simulation of SEIR model for *α* = 0.7.

In this example, some parameters of the model are chosen such that 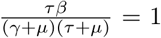 which can be observed from the equation (39).

## 4 Numerical solution of the model with Mittag-Leffler process

In this section, we adopt the numerical scheme for the model where first part is classical and second part is the fractional derivative with the generalized Mittag-Leffler kernel. It is important noting that a model with the generalized Mittag-Leffler helps capture processes,with a passage from streched exponential to power-law with no steady state.

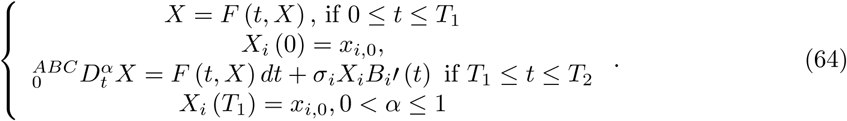

Applying the Atangana-Baleanu integral, we obtain the following equality

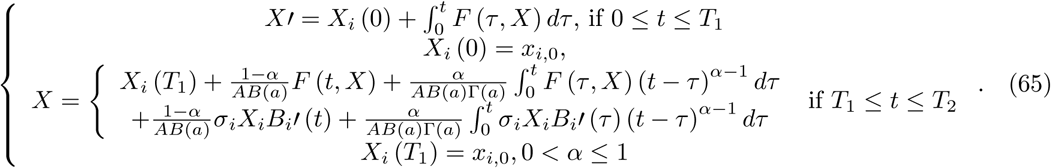

We divide [0, *T*] in three

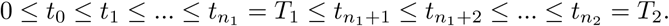

Replacing the functions *f*_*i*_ (*t, X*_*i*_) by their the Newton polynomial within [*t*_*n*_, *t*_*n*+1_], the following numerical scheme is obtained

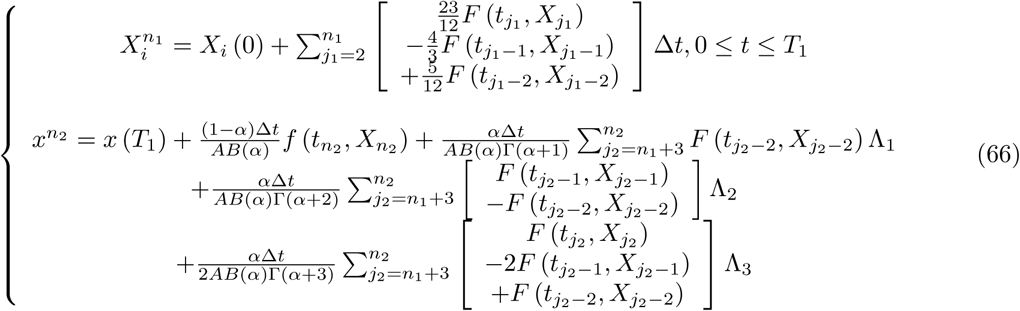

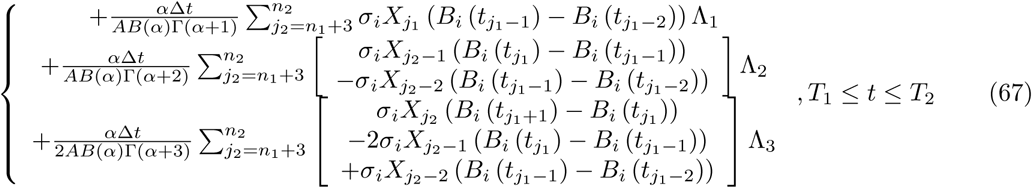

where

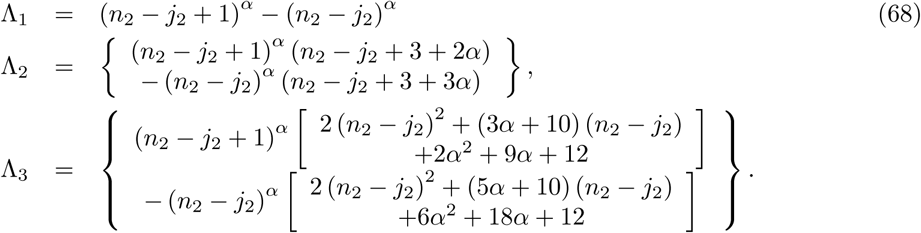

For this model, the parameters and initial conditions are considered as

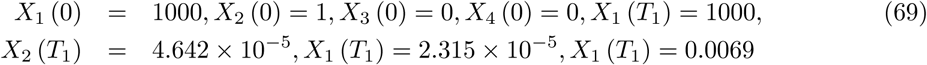

and

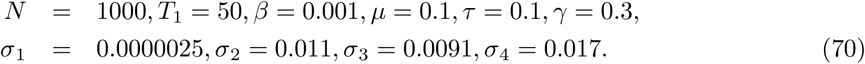

The simulation of the numerical solution of the model is performed in Figure 3.

**Figure 3.**
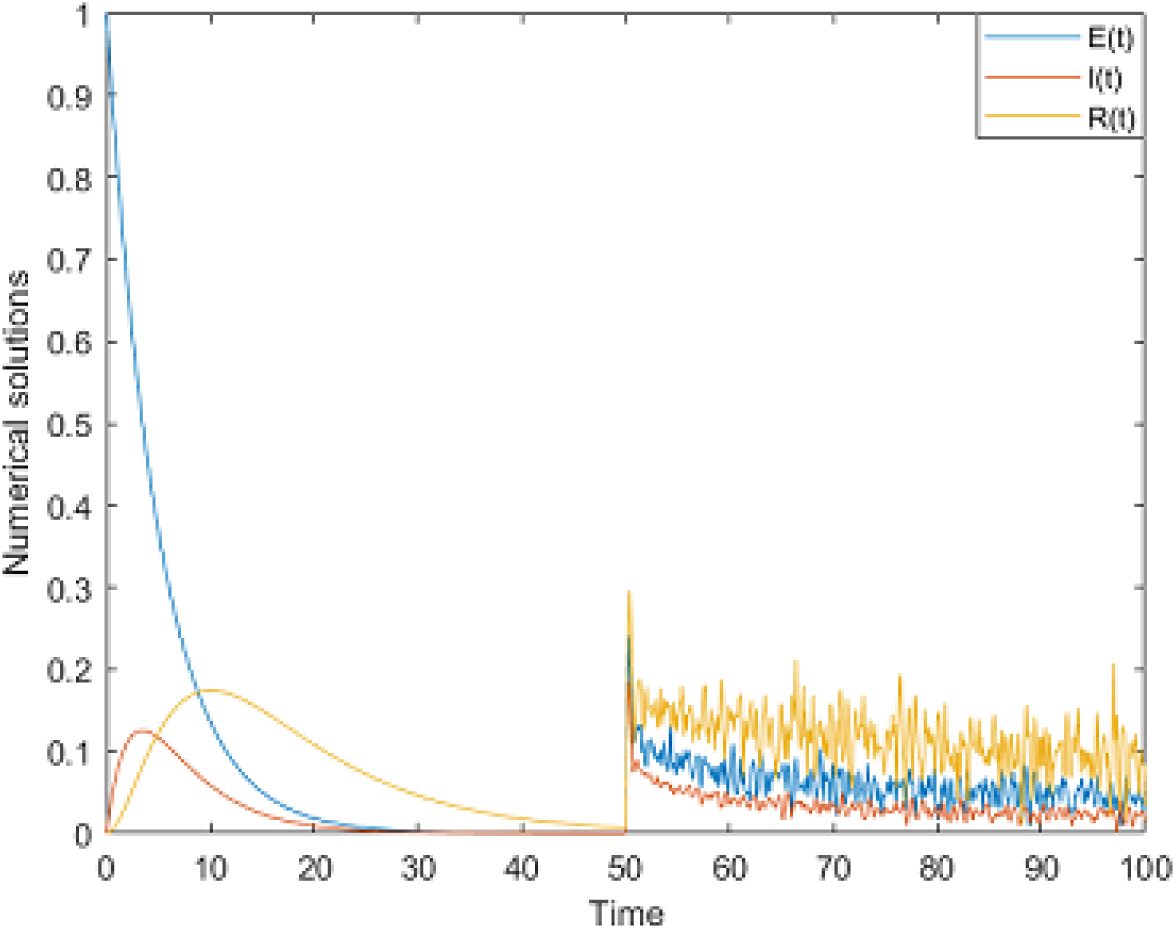
Numerical simulation of SEIR model for *α* = 0.6.

In this example, the parameters of the model are chosen such that 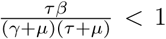 which is seen from the equation (39).

## 5 Numerical solution of the model with power-law process

In this section, we consider the following model

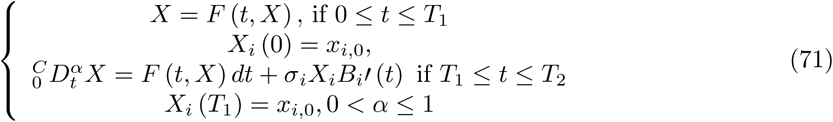

where first part is classical and second part is the fractional derivative with the generalized Mittag-Leffler kernel. Applying the associated integral, we obtain the following equality

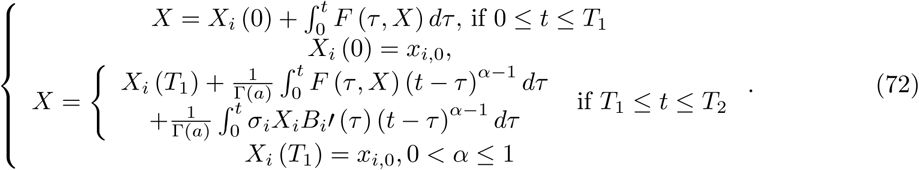

We divide [0, *T*] in three

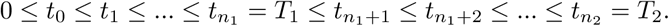

Replacing the functions *f*_*i*_ (*t, X*_*i*_) by their the Newton polynomial within [*t*_*n*_, *t*_*n*+1_], the associated model can be solved by the following algorithm

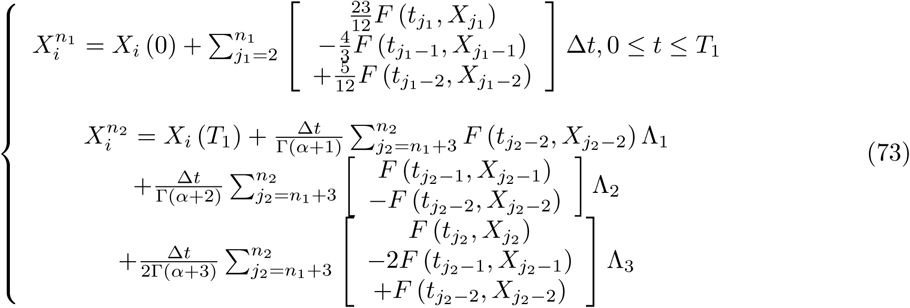

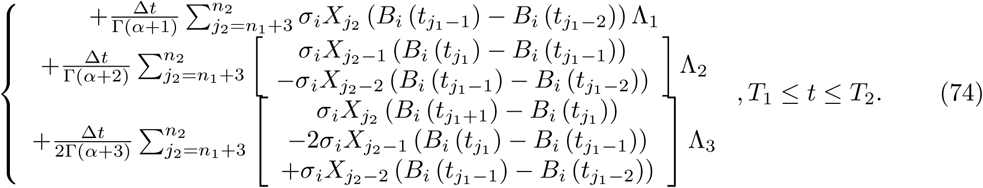

The parameters and initial conditions are given as

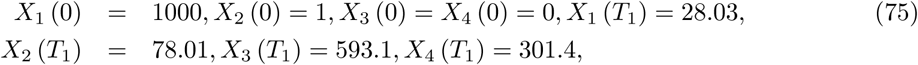

and

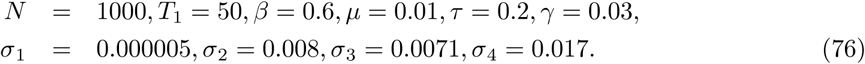

The simulation of the numerical solution of the model is presented in Figure 4.

**Figure 4.**
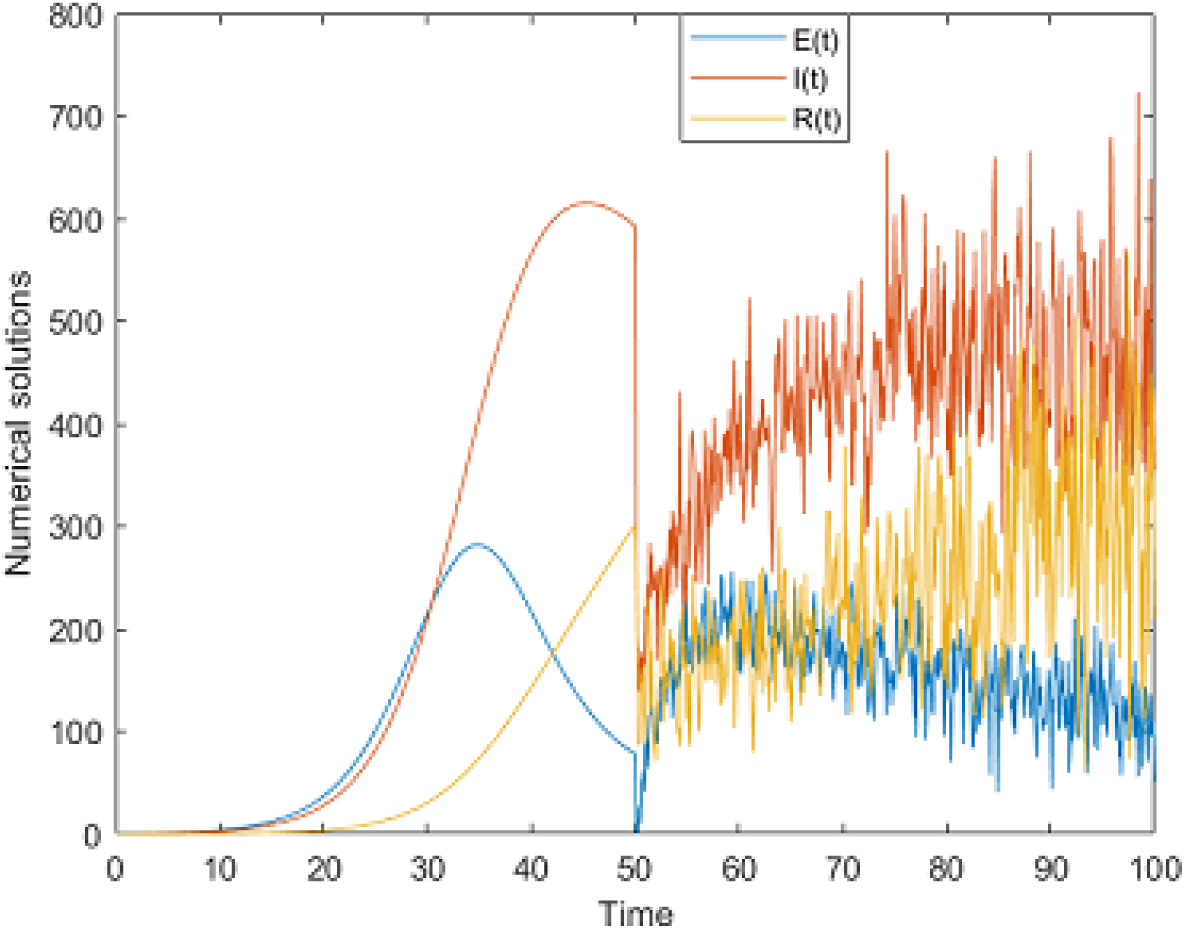
Numerical simulation of SEIR model for *α* = 0.75.

In this example, the parameters of the model are chosen such that 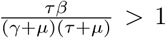 which is seen from the equation (39).

## 6 Conclusion

In classical calculus, analysis of the first derivative of a given function helps to understand the change in such a function. On the other hand, this formula is used to calculate the velocity of a moving object. However, analysis using first derivative does not always a priori provide clear indication variation of function, therefore, an analysis of second derivative is required. The analysis of the second derivative provides information of inflection points, local maximum and minimum. These elementary analyses can be used to better understand spread patterns in epidemiological modelling. A new concept was recently introduced and named Strength number that is obtained by considering the second derivative of the nonlinear part of a given model of an infectious disease, then, the next generation matrix methodology is used to obtain the strength number. It was argued that such numbers could help detect waves or instability in a model. In this work, such a concept is applied together with second derivative analysis in a simple SEIR problem. The obtained strength number was negative, one equilibrium point for the model with second derivatives, a clear indication that such a model could have only one wave then die out. We have used piecewise differential operators to add stochastic behavior in the model.

## Data Availability

Not applicable

